# Seroprevalence of anti-SARS-COV-2 antibodies in blood donors from Nuevo Leon state, Mexico, during the beginning of the COVID-19 pandemic

**DOI:** 10.1101/2020.11.28.20240325

**Authors:** Natalia Martinez-Acuña, Diana Avalos-Nolazco, Diana Rodriguez-Rodriguez, Cynthia Martinez-Liu, Rogelio Cazares Taméz, Amador Flores-Arechiga, Fernando Perez-Chavez, Sergio Ayala-de-la-Cruz, Eduardo Cienfuegos-Pecina, Erik Diaz-Chuc, Daniel Arellanos-Soto, Gerardo Padilla-Rivas, Javier Ramos-Jimenez, Kame Galan-Huerta, Sonia Lozano-Sepulveda, Elvira Garza-González, Consuelo Treviño-Garza, Roberto Montes-de-Oca-Luna, Aurora Beatriz Lee, Manuel de-la-O-Cavazos, Ana Maria Rivas-Estilla

## Abstract

**Background:** The progression and distribution of SARS-CoV-2 is unknown because typically only symptomatic individuals are diagnosed.

**Aim:** We evaluated the seroprevalence of anti-SARS-CoV-2 in blood donors in Nuevo Leon, Mexico as a strategy for asymptomatic case detection of COVID-19 and epidemic progression.

**Methods/Materials:** We tested 1968 blood donors that attended two regional donation centers in Northeast Mexico from January 1^st^ to August 30, 2020, to identify anti-SARS-CoV-2 IgG by chemiluminescent immunoassay. Additionally, routine tests for donors including Brucella, Chagas, HCV, VDRL, HIV-1, and HBsAg identification were performed.

**Results:** We found 77 donors reactive for anti-SARS-CoV-2 IgG (seroprevalence 3.99%) and none of them had reported recent COVID-19 symptoms. Donors aged 18 to 49 years (89.5%) were more likely to be seropositive compared to those aged 50 years or older (10.5%) (P<0.001). Prevalence of antibodies increased each epidemiological (EPI) week, parallel to the report of confirmed cases by RT-PCR, identifying the highest prevalence between EPI week 33 and 35 (10.2% to 19%). The metropolitan area of Monterrey recorded the highest number of cases. Routine tests showed that the prevalence of anti-Brucella was 0.13%, anti-HCV 0.5%, anti-HIV-1-2 0.14%, HBsAg 0.16%, Chagas 0.48% and syphilis 1.06%.

**Conclusions:** There is a growing trend of seroprevalence over time, parallel to the constantly increasing epidemic curve in our region and it was higher under 49 years of age associated with asymptomatic infection in donors from the Nuevo Leon area. Detection of anti-SARS-CoV-2 in blood donors is a potential tool for tracking the progression and identifying population exposure during the SARS-CoV-2 outbreak.

## Introduction

Severe acute respiratory syndrome coronavirus 2 (SARS-COV-2) producing coronavirus disease 2019 (COVID-19) has spread around the world becoming a major public health problem infecting over 46 million individuals worldwide leading to over 1 million deaths (Johns Hopkins University). In Mexico, official statistics report around 1,078,594 cases and 104,242 deaths, while Nuevo Leon state has recorded 99,376 cases with 5,419 deaths. Mortality in Mexico reaches 9.7% (Health Secretariat, Mexico, November 27^th^,2020, https://www.gob.mx/salud).

The clinical presentation in COVID-19 patients includes fever, dry cough, and fatigue as the most common symptoms that may appear 3 - 14 days after virus infection. Several reports have suggested that SARS-CoV-2 infections can evolve as asymptomatic cases, where no or non-perceptible clinical manifestations occur. It is reported that the asymptomatic incidence can range from 1.2 to 12.9% in large populations. However, other studies with a smaller sample size reported a much higher proportion reaching up to 87.9% [1]. Reports have demonstrated that viral loads are very similar in symptomatic and asymptomatic groups making the latter capable of spreading SARS-CoV-2 despite the absence of clinical manifestations [2,3]. Most governments have not officially established screening to identify asymptomatic cases, as a strategy for the detection of infected cases, mainly because there still is not a clear indicator of infection that serves as a reference for molecular screening. More detailed epidemiologic data and the recalculation of prevalence and fatality rates of the COVID-19 pandemic are necessary to consider the real number of both symptomatic and asymptomatic infected subjects.

Specific antibodies against SARS-CoV-2 appear between 4 and 5 days (IgM antibodies) after infection and most of the patients seroconvert (IgG antibodies) within the first 3 weeks after the infection is established reaching detectable levels in most infected individuals 15 days following the onset of COVID-19 symptoms [4–6]. Robust titers of specific neutralizing antibodies can be detected for at least 5 months post-SARS-CoV-2 infection [7].

While most cases of COVID-19 have mild-to-moderate symptoms, the antibody responses in less severe infections are not well characterized. Estimating the seroprevalence of anti-SARS-CoV-2 in blood donors is a powerful and cost-effective strategy to monitor virus circulation among healthy people. Early studies focusing on blood donors to identify COVID-19 infected persons were conducted in Denmark and the Netherlands [8,9]. The seroprevalence of SARS-CoV-2 antibodies determined using this approach varies from 0.1% (San Francisco Bay Area) [10] to 5.6% (Kenya) but can reach values of up to 9% when they are adjusted by a determined geographical region [11].

To date, few data have documented the prevalence of SARS-CoV-2-antibodies in the general population and asymptomatic outpatients in Mexico. In this work, we conducted a seroprevalence survey using residual plasma samples to identify SARS-CoV-2 specific IgG antibodies among blood donors of two donation venues in Nuevo Leon, Mexico from January to the end of August 2020, which include pre-pandemic and ongoing pandemic timing in our country. Identification of asymptomatic cases of COVID-19 allowed us to add information to track the progression of the pandemic in our country.

## Material and Methods

### Study design and subjects

We studied samples from blood donors living in Nuevo Leon state (which is located in northeast Mexico and borders with the state of Texas, United States) who attended two donation venues, the Blood Bank Center at the “Dr. Jose E. Gonzalez” University Hospital and the Blood Bank of the Transfusion Center of the Secretariat of Health of Nuevo Leon state, Mexico. We conducted a cross-sectional seroprevalence survey of IgG antibodies from January 1^st^ to August 30^th^, 2020 (pre-pandemic and the start of the pandemic phase). Consecutively, a non-selected subset of 1968 blood donors (ages 18–65) was selected according to the requirements of the Mexican Official Norm NOM-253-SSA1-2012.

Donors were asked whether they experienced symptoms compatible with the presence of a viral illness during the previous three weeks. To confirm the status of asymptomatic, all donors underwent a medical interview and had no physically detectable symptoms of infection. We used ArcGIS v10.2.2 (ESRI, Redlands, CA, USA) to indicate the municipality of donors actual residence. The research protocol was reviewed and approved by the institutional review board. We used exclusively residual plasma or serum from routine laboratory diagnosis of blood donors; therefore, the need for informed consent was waived. Data regarding demographic and clinical epidemiological characteristics were collected from the anonymous archive and donor management software of the Blood Bank through a standardized questionnaire.

### Serological analysis

Plasma samples were obtained from each donor to perform serological tests. Qualitative detection of anti-SARS-CoV-2 IgG against the nucleocapsid protein was performed according to the manufacture’s protocol using a chemiluminescent microparticle immunoassay detection kit (SARS-CoV-2 IgG ARCHITECT; Abbott Laboratories; reference 06R8620) in residual plasma specimens. In this study, an index S/C threshold of 1.5 or superior was taken as a positive result. We performed routine serologic tests for blood donors which include antibodies detection against *Treponema pallidum* (VDRL), Hepatitis C virus (HCV), human immunodeficiency virus 1/2 (HIV-1/2), Hepatitis B Virus (HBV), and *Trypanosoma cruzi* (Chagas disease), using an Architect i2000 SR analyzer (Abbott Diagnostics, Chicago, USA). Detection of *Brucella spp*, was performed using the Rose Bengal Test (Licon, Mexico City).

### Statistical Analysis and Ethical statement

Prevalence was calculated for the total analyzed study population and by identified strata and analyzed by the Chi-squared test. All data were entered into a Microsoft Access database (Microsoft Access 2002, Microsoft Corp., Redmond, WA) and exported to SPSS (version 13.0 for Windows; SPSS Inc., Chicago, IL) for statistical analyses. We used exclusively residual plasma or serum from routine laboratory diagnosis of blood donors; therefore, the need for informed consent was waived. The donation was made under National and International Blood Establishment Authorization.

## Results

### Study population

A total of 1968 residual plasma specimens were analyzed for the presence of anti-SARS-CoV-2 IgG during the period between January 1^st^ to August 30^th^, 2020 (Fig. 1). The studied period comprises the pre-pandemic phase in our country (with 37 samples obtained from epidemiological (EPI) week 1 to 10) and the start of the sanitary contingency phase referred to as the pandemic group which was used to perform the statistical analysis (with 1931 samples from EPI week 11 to 36) (Fig. 1A and B). None of the seropositive blood donors reported a known positive medical history of COVID-19 before donation.

**Figure 1.**
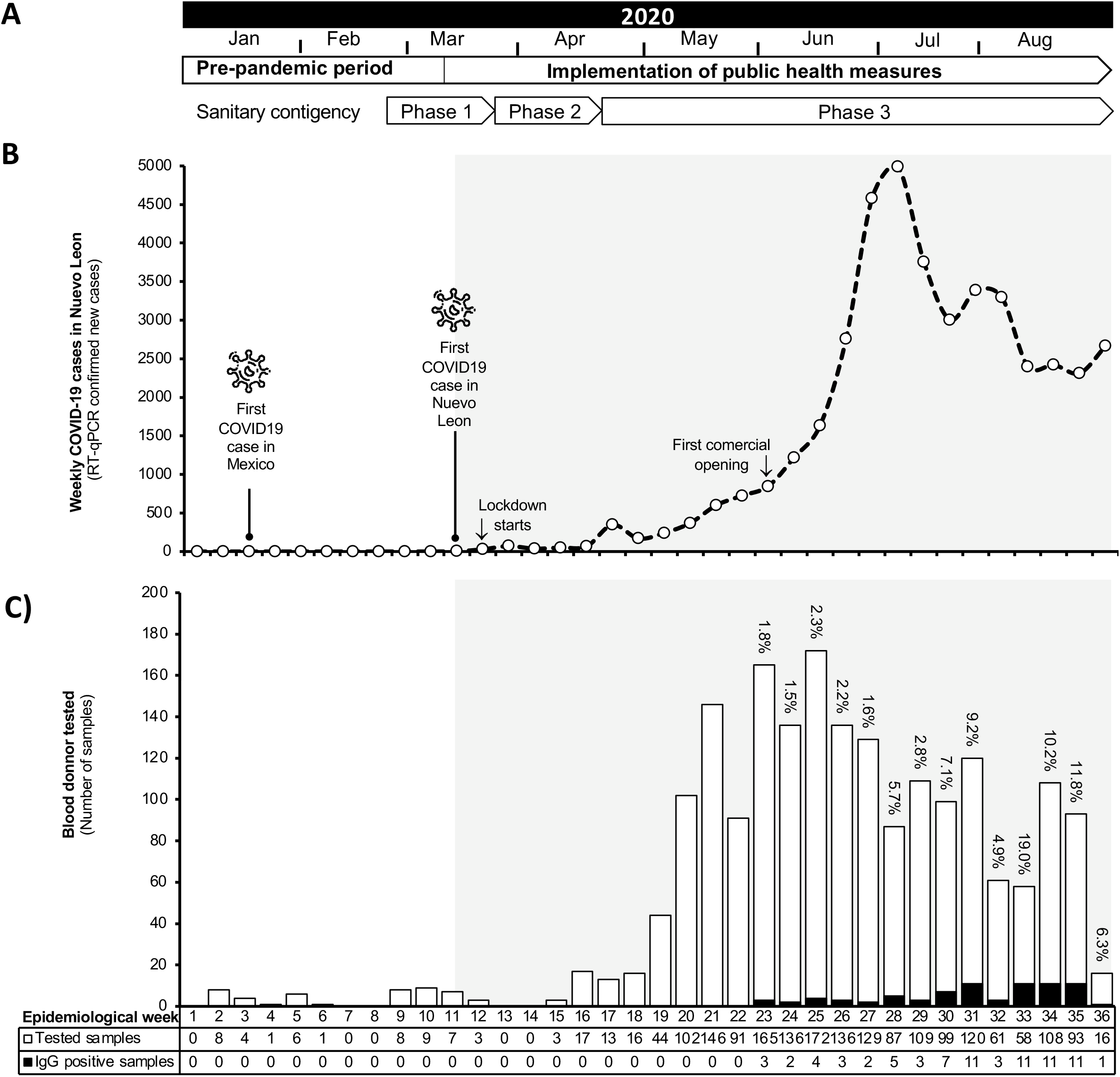
Weekly seroprevalence of SARS-CoV-2 antibody during the pandemic walkthrough in Mexico from January to August 2020 in Nuevo Leon, Mexico. A) Time distribution of the observed period. Relevant events to the SARS-CoV-2 pandemic in Mexico and social distancing measurements are indicated. B) Curve of RT-qPCR confirmed new COVID-19 cases per epidemiological week recorded in the State of Nuevo Leon. The gray shadow indicates the pandemic period. Regional relevant events are indicated. C) A total of 1968 plasma samples were collected from January 7 to August 31 of 2020 in two regional blood bank units. Plasma was analyzed using an anti-SARS-COV-2 IgG detection kit from Abbott Biotechnology. Cutoff values of 1.5 were considered positive. The data were grouped per EPI week and prevalence was estimated by dividing the number of positive cases (black bars) by the number of tested samples (white bars) per each week. Weekly seroprevalence of SARS-CoV-2 IgG is shown on the top of each bar. The gray shadow indicates the pandemic period.

### Detection of anti-SARS-CoV-2 IgG in plasma derived from blood donors

We detected 77 reactive donors among the pandemic group giving an overall seroprevalence of anti-SARS-CoV-2 IgG of 3.99% (77/1931), from which 41 were male (53.2%) and 36 were female (46.7%). Specific seroprevalence per epidemiological week (EPI week) is reported in figure 1B. At this point of the pandemic, there was an increasing trend on the number of qRT-PCR confirmed and the suspected cases in the state, which was also parallel to the detection of the first donor detected on EPI week 23 (Fig. 1B and C).

The main characteristics and epidemiological data of the studied donors and specific prevalence calculated for sex, age group, blood type, and educational level are shown in Table 1. The mean age for blood donors was 34.8 years (range 18 to 65 years). Most participants were men (N=1436, 74.5%). Donors aged 18-49 years (89.5%) were more likely to be seropositive compared to those age 50 years or older (10.5%) (P<0.001). Regarding each group, the highest prevalence was observed among female donors (7.27%, *p* < 0.001), persons with a higher education level (8.6%, *p* < 0.001), and a BMI above 35 (5.33 %, *p*=0.566). Individuals with blood types B (5.18 %) and AB (6.25 %) were more likely to test positive. Two donors that assisted in July reported developing fever two days after donation and were confirmed as positive for SARS-CoV-2 by RT-qPCR nasopharyngeal detection but were negative for antibody testing.

**Table 1.** Blood donor’s characteristics and crude prevalence of antibodies against SARS-CoV-2 observed in collected samples from Nuevo Leon State, Mexico, during the pandemic period (EPI week 11 to 36, 2020).

| Variable | Group | Total samples (n=1931) | Positive cases (n = 77) | Prevalence positive cases (%) | Prevalence per group (3.99%) | p-value* |
| --- | --- | --- | --- | --- | --- | --- |
| **Sex** | M | 1436 | 41 | (53.2 %) | 2.86 % | <0.0001 |
|  | F | 495 | 36 | (46.8 %) | 7.27 % |  |
| **Age group** | 18-29 | 695 | 29 | (37.7 %) | 4.17 % | 0.9063 |
|  | 30-49 | 1050 | 40 | (51.9 %) | 3.80 % |  |
|  | 50-65 | 186 | 8 | (10.4 %) | 4.30 % |  |
| **ABO blood type** | A | 474 | 21 | (27.3 %) | 4.43 % | 0.7303 |
|  | B | 135 | 7 | (9.1 %) | 5.18 % |  |
|  | AB | 16 | 1 | (1.3 %) | 6.25 % |  |
|  | O | 1306 | 48 | (62.3 %) | 3.67 % |  |
| **RH Blood type** | + | 1849 | 73 | (94.8 %) | 3.94 % | 0.5660 |
|  | - | 82 | 4 | (5.2 %) | 4.87 % |  |
| **BMI** | <18.5 | 2 | 0 | (0.0 %) | 0% | 0.5135 |
|  | 18.5-25 | 424 | 14 | (18.2 %) | 3.30 % |  |
|  | 25-30 | 834 | 29 | (37.7 %) | 3.47 % |  |
|  | 30-35 | 465 | 23 | (29.9 %) | 4.94 % |  |
|  | >35 | 206 | 11 | (14.3 %) | 5.33 % |  |
| **Educational level** | Basic | 913 | 8 | (10.4 %) | 0.87% | <0.0001 |
|  | Middle | 588 | 32 | (41.5 %) | 5.44% |  |
|  | Higher | 430 | 37 | (48.1 %) | 8.60% |  |

\*Chi-square test

During the pre-pandemic weeks, SARS-CoV-2 antibodies were not detected in any tested sample (Fig. 1A and B). The first positive plasma for IgG antibodies in donors was detected until May 31^st^ (EPI week 23), twelve weeks upon the first confirmed case was reported in Nuevo Leon (Fig. 1B and C). During the study, there was an increasing trend over time in weekly IgG antibody seroprevalence (the number of positive cases/samples collected per week) from 1.8% in May until a maximum value of 19% by the end of August 2020 (EPI week 33) (Fig. 1C).

### Geographical distribution of observed asymptomatic cases of SARS-CoV-2 infections

Reactive donors for anti-SARS-CoV-2 were concentrated in the major urban area. Collected plasma samples were from 40/51 municipalities of Nuevo Leon state. The SARS-CoV-2 IgG positive cases were distributed across 19 of Nuevo Leon’s municipalities and were frequently observed in the Monterrey metropolitan area, also including Apodaca, General Escobedo, and Benito Juarez municipalities (Figure 2A). Although the major proportion of cases relative to analyzed samples was associated with the rural area: Cienega de Flores (2/6, 33%), Allende (2/14, 14%), and Pesqueria (2/25, 8%). Inside the Metropolitan area, the proportion of cases varied from 1.2% (1/86) in Santa Catarina to 5.7% (10/175) in Apodaca (Figure 2B and C).

**Figure 2.**
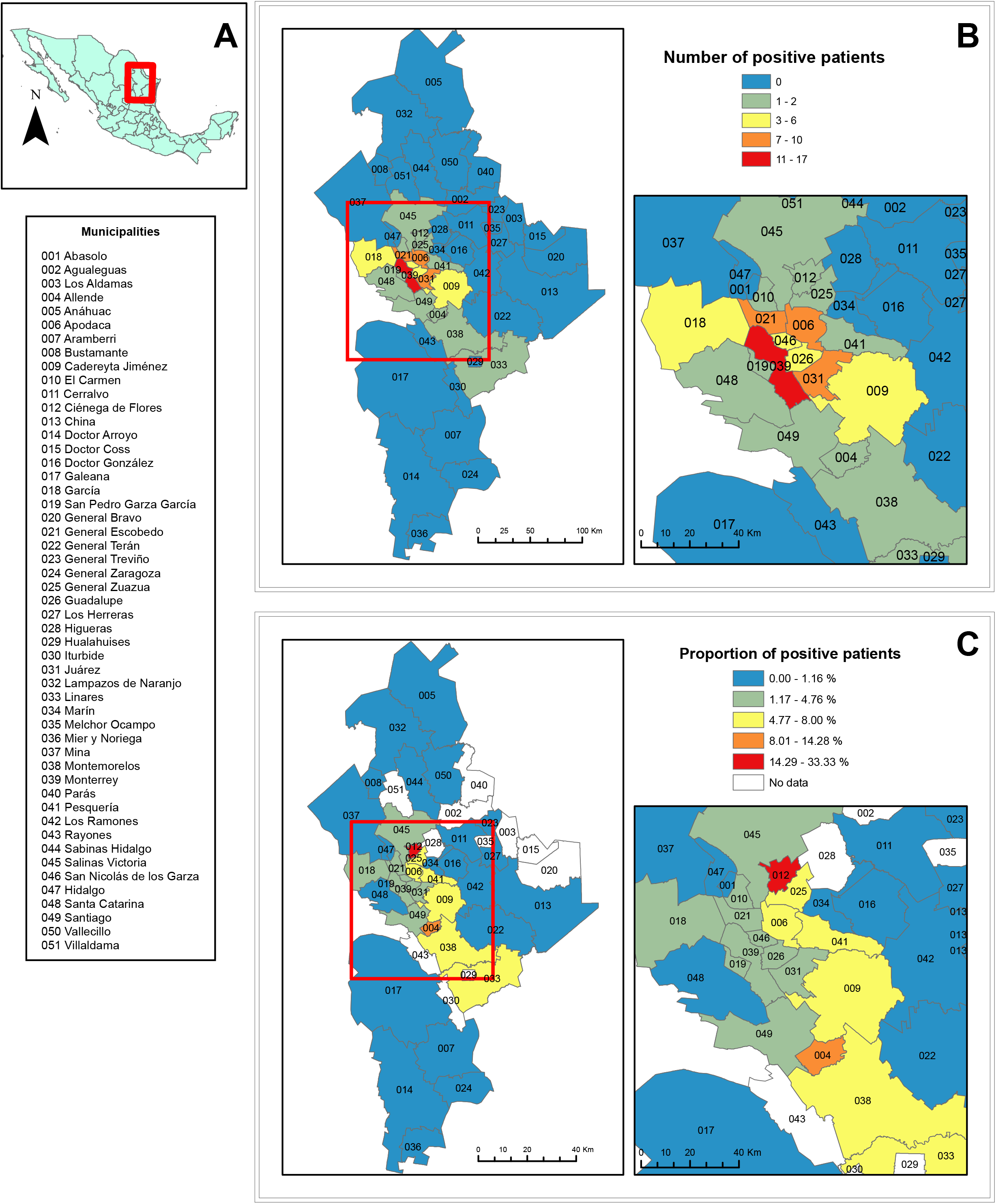
Geographical distribution of total asymptomatic SARS-CoV-2 infection events determined by SARS-CoV-2 antibody IgG detection in the Nuevo Leon Area.. A) Studied Municipalities of Nuevo Leon state; B) Number of positive cases of COVID-19 by municipality; C) Proportion of positive cases of COVID-19 by municipality.

### SARS-COV-2 prevalence and other relevant infectious diseases in blood donors

The results of routine tests in the blood donors tested showed that the prevalence of *Brucella spp* Abs was 0.13%, anti-HCV was 0.5%, anti-HIV-1-2 was 0.14%, HBsAg was 0.16%, *T. cruzi* was 0.48%, and *T. pallidum* with 1.06%; while Anti-SARS-CoV-2 IgG seroprevalence was much higher (average 3.99%) (Figure 3). These values seem to be similar to previous reports in our region[12,13].

**Figure 3.**
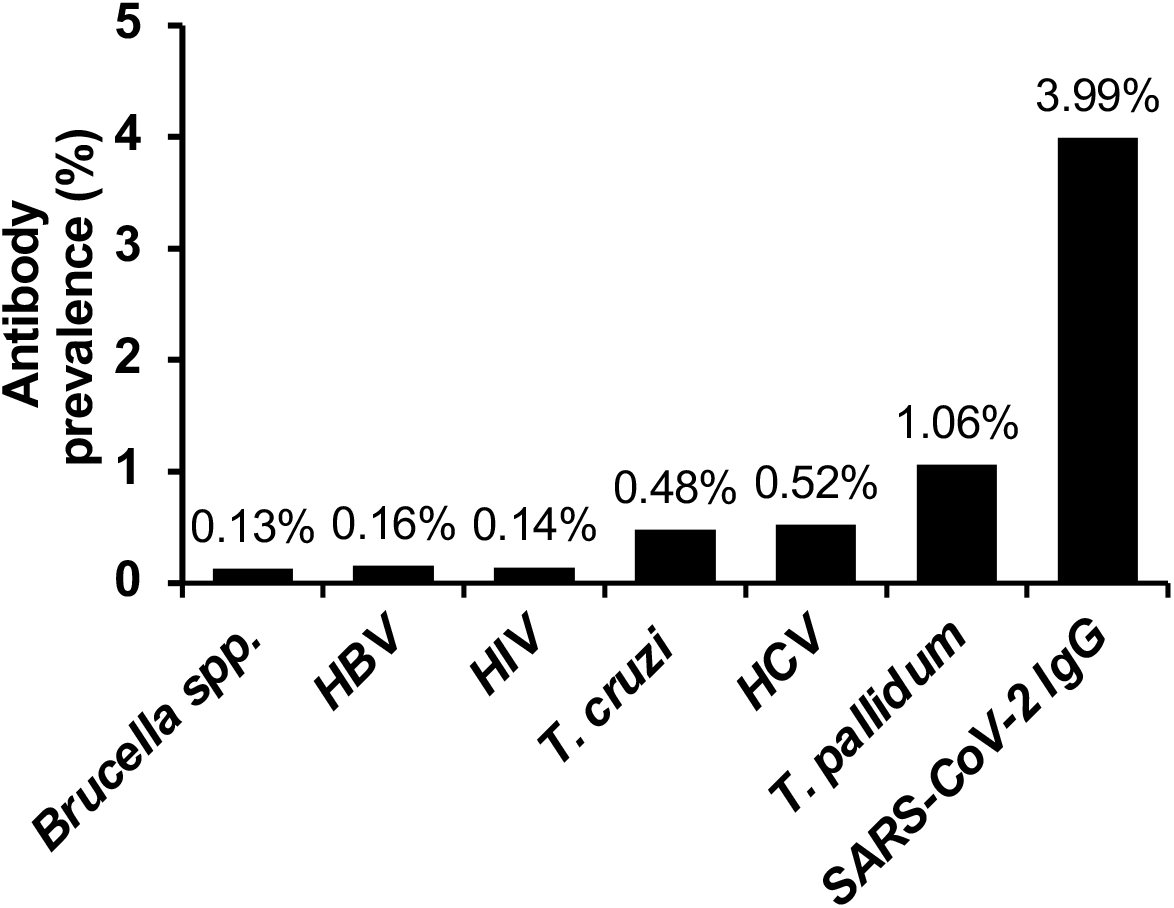
Prevalence of SARS-COV-2 IgG antibodies and other blood-borne infectious diseases detected in studied donors from January to August 2020.

## Discussion

While RT-PCR assays are world widely used to confirm COVID-19, serological testing offers a means of identifying individuals who previously experienced asymptomatic infections, as well as those who presented symptomatic infections but have resolved the infection.

In this study, we identified that in the beginning stage of the COVID-19 pandemic in Mexico, 3.9% of the evaluated donors presented positive IgG antibodies for SARS-CoV-2, which represents a much lower percentage of symptomatic infected individuals detected by RT-qPCR throughout the country. This is due in part to the fact that at the government level, the search strategy for infected cases focuses primarily on the study of symptomatic people. These strategies will change as the pandemic progresses and the number of infected asymptomatic subjects becomes more evident, whose role becomes very important in the transmission and control of the disease.

Regarding the overall seroprevalence data reported here (3.9%), we found a higher prevalence than that reported in Wuhan, China with 2.29% [14], the Netherlands with 2.7%[9], and Denmark with 1.7% among blood donors [8]. In a study performed by the American Red Cross in the United States, all blood donations from June to August 2020 were tested for anti-SARS-CoV-2 IgG antibodies. They found that 1.82%. (17,336/953,926) of the donations were positive for SARS-CoV-2 IgG antibodies [15]. We also found, as in other regions, that anti-SARS-CoV-2 seroprevalence in blood donors was starting slowly and then growing rapidly and varied by regions, as demonstrated by a report from the United Kingdom where they evaluated the progression and geographical distribution of SARS-CoV-2 neutralizing antibodies [16]. Similarly, analysis of the distribution of samples showed that most of these samples were in urban areas. As we described before, seroprevalence through EPI weeks was dynamic increasing from 1.8% in May to 19% in August 2020 and seroprevalence increased significantly over time across all regions. This increase may be due to donors with higher rates of prior exposure and may also reflect increased exposure in the general population.

Today, there are still few studies on the prevalence of SARS-CoV-2 in blood donors. With the information available worldwide so far, it has been estimated that 15% to 46% of SARS-CoV-2 infections are asymptomatic [17] but it can vary widely according to the time of exposure of the population, the epidemiological management of the pandemic, and other factors that are still unknown that affect different regions in the world.

As limitations of our study, first, we note that the seroprevalence estimated from this study may not reflect the true underlying proportion exposed to SARS-CoV-2 in our country because blood donors are not representative of the overall population; second, we did not test samples for virus neutralization and therefore the neutralizing activities of the detected IgG antibodies are unknown. Third, a rapid decline in antibody titers and pro-inflammatory cytokines may be a common feature of non-severe SARS-CoV-2 infection. It has been suggested that asymptomatic individuals may have a weaker immune response to SARS-CoV-2 infection in contrast to symptomatic [18]. At the time of this study, SARS-CoV-2 serological tests available only allow IgG analysis and we only observed past infections with at least two or three weeks of evolution; more recent infectious events may be unnoticed. This implies that the cases may be higher. It is important to consider that the apparent decrease in the titers may lead to false-negative results over subsequent months, a phenomenon not yet well determined. In this study, samples were collected during the 5 months after diagnosis of the first confirmed case by RT-qPCR in Nuevo Leon. For this reason, this study is unlikely to be hampered by a drop in SARS-CoV-2 antibody levels. On the other hand, it is important to consider that for future seroprevalence studies it will be important to carry out more studies to validate the lifespan of the antibodies and consider a possible underestimate of the real exposure of the population.

Also, the results of this study allow us to highlight two important points: there is a growing trend of seroprevalence over time, parallel to the constantly increasing epidemic curve in our region, and the higher prevalence found in subjects under 49 years of age is associated with asymptomatic infection of these donors. Nevertheless, we demonstrated that screening blood donors for SARS-CoV-2 antibodies strengthens existing evidence that this group can be used as a sentinel population to track the progression of the COVID-19 pandemic.

## Data Availability

All data referred in this manuscript are available

## Acknowledgments

We thank the staff of the Central Blood Bank of the Department of Clinical Pathology, “Dr. Jose E. Gonzalez” University Hospital and School of Medicine, UANL; and Secretaria de Salud de Nuevo Leon (Transfusion Center CETS) for their technical help and facilitating this study. This work was supported in part by CONACYT Grants: 0312135 to NMA; 314867 to AMRE and 312368 to SALS.

